# Reframing infectious diseases and antimicrobial resistance through a One Health lens: A systems approach to understanding complexity

**DOI:** 10.1101/2025.08.01.25332650

**Authors:** Yen Pham, Sonja Janson, Teresa M. Wozniak

**Affiliations:** Australian e-Health Research Centre, Commonwealth Scientific and Industrial Research Organisation, Brisbane, Queensland, Australia; Department of Infectious Diseases, Royal Darwin Hospital, Darwin, Northern Territory, Australia; James Cook University, Queensland, Australia

## Abstract

**Background:** Antimicrobial resistance (AMR) is a complex global health threat with uneven impacts across regions. In Australia, the AMR burden varies geographically, with up to 50% of infections now resistant to antibiotics in northern Australia.

**Methods:** This qualitative study engaged stakeholders across One Health sectors in the Northern Territory to explore perspectives on AMR. Thematic analysis of interview data informed the development of a causal loop diagram, grounded in systems thinking, to illustrate interactions between factors driving AMR. This paper focuses on the key factors and feedback loops influencing AMR in human health.

**Findings:** Thirty-five participants working across human, animal, environmental and cross-sectoral health systems were engaged through interviews and focus groups. The resulting causal loop diagram revealed dynamic, interconnected factors contributing to AMR through reinforcing and balancing feedback loops. Key drivers included antibiotic prescribing and use in both human and animal health, as well as antimicrobial residues in the environment. However, broader structural determinants—such as geographical isolation, health system limitations and social inequities—also shaped AMR dynamics. Therefore, reducing antibiotic use alone is unlikely to sufficiently control AMR. While antimicrobial stewardship and infection control remain critical, additional leverage points for interventions were identified. These include the promotion of culturally safe care, healthcare workforce training and retention, improved AMR awareness among professionals and communities, and investment in upstream social determinants of health, such as functional housing and access to essential health infrastructure.

**Conclusions:** In regions with high infectious disease burdens, addressing AMR requires a shift from reactive, clinical models to proactive, community-based strategies. Given its systemic complexity, AMR cannot be addressed by health systems alone or through siloed interventions. A coordinated, multi-sectoral and systems-informed approach is essential for sustainable impact.

## 1. Introduction

Antimicrobial resistance (AMR) is a critical global health concern, contributing to approximately 4.95 million deaths in 2019 [1] with significant regional disparities. By 2050, annual global deaths linked to AMR are projected to reach 8.22 million [2]. In Australia, the burden is disproportionately high in regional and remote settings of northern Australia, where the risk of infectious diseases is three times greater than in urban areas, and up to 50% of *Staphylococcus aureus* infections are methicillin-resistant [3, 4]. In these contexts, infectious diseases not only strain healthcare systems but also undermine community wellbeing and socioeconomic stability.

Although AMR is often framed within a clinical context, the emergence and spread of resistant pathogens extend beyond human health, reflecting a complex network of interconnected drivers across the One Health spectrum [5]. For example, antimicrobials and resistant microorganisms excreted by humans or animals can enter the environment through inadequately treated waste [6]. In addition, anthropogenic activities, such as the discharge of heavy metals [7], irrigation with untreated sewage, and application of livestock manure [8] exert selective pressure on microbial populations. This, in turn, facilitates the horizontal transfer of resistant genes, expanding the environmental reservoir of resistance and increasing the risk of transmission to humans and animals [9]. Despite the dynamic complexity of these One Health interrelationships, AMR is often addressed in disciplinary silos, limiting our capacity to understand and develop effective, coordinated responses [10].

Systems thinking provides a valuable framework for understanding how various factors within complex systems are interrelated [11]. It offers tools and methods to analyse these interconnections and design long-lasting interventions for complex challenges [12]. This approach is particularly well-suited to addressing the multifaceted factors driving AMR across One Health sectors and identifying leverage points that enable sustainable, systemic change [5, 13]. However, a recent systematic review has revealed that systems thinking approaches remain underutilised in AMR research, particularly within the One Health context [10], despite their demonstrated success in other domains. While there is broad consensus that AMR should be addressed through a One Health lens, gaps remain in our understanding of the dynamic relationships and feedback processes involved, hindering the ability to develop effective responses.

This study applies system dynamics, a systems thinking approach, to investigate the interconnected factors and feedback mechanisms influencing AMR in the Northern Territory of Australia, a region with high infectious disease burdens and exceedingly high AMR rates. By addressing key knowledge gaps in understanding AMR dynamics across One Health domains, this research aims to support the development of coordinated, comprehensive and sustainable strategies to mitigate AMR. This paper presents findings focused on the human health dimensions of AMR, and forms part of a larger body of work examining its evolution and spread across all One Health sectors.

## 2. Material and methods

### 2.1. Causal loop diagrams

This study developed a causal loop diagram (CLD), a qualitative modelling tool that visualises the feedback structures underlying complex problems—a crucial step in the system dynamics process [14]. Drawing on stakeholder input, the study aimed to foster a shared understanding among stakeholders of AMR evolution and spread by exploring the system-wide dynamics influencing AMR across the One Health spectrum and identifying potential leverage points for interventions.

Causal loop models or CLDs consist of variables connected by arrows that indicate causal relationships. If an increase (or decrease) in variable A (e.g., infections) leads to a corresponding increase (or decrease) in variable B (e.g., antibiotic use), a *‘+’* symbol is used to indicate a *positive relationship*. Conversely, if an increase in A (e.g., infection control) leads to a decrease in B (e.g., infections), a *‘-‘* symbol represents a *negative relationship* [12]. CLDs feature two types of feedback loops that influence system dynamics or behaviour over time: *reinforcing (R) or positive loops*, which amplify change, and *balancing (B) or negative loops*, which resist change and promote system stability or equilibrium [12]. *‘Delays’ (//)* indicate time lags between causes and effects, often contributing to trade-offs between short- and long-term outcomes, and may lead to unintended consequences [14]. CLDs allow for the identification of ‘*leverage points*’—specific areas in a system where small shifts can produce significant, lasting impact [12].

### 2.2. Data collection and analysis

Data for the CLD were collected through stakeholder interviews and focus groups. The study followed the Consolidated Criteria for Reporting Qualitative Research checklist [15]. Ethical approval was obtained from the Human Research Ethics Committee of the Commonwealth Scientific and Industrial Research Organisation, as well as from the NT Health and Menzies School of Health Research. All participants provided either written or verbal consent at the start of recorded interviews or focus groups. Participant recruitment took place between 23 August 2023 and 30 December 2024.

A systemic approach to stakeholder identification was adapted from Schiller, Winters [16], involving the development of a stakeholder map with key categories and groups associated with AMR. Participants were initially recruited through the investigators’ professional networks using purposive sampling [17]. At the end of each interview, participants were invited to share recruitment materials or refer others (snowball sampling as described in Kirchherr and Charles [18]).

Semi-structured interviews, widely recognised as effective for model formulation [14], were used to explore both root causes and causal relationships in the AMR system. Questions were tailored to participants’ professional background. Interview data informed the development of a preliminary CLD, which was then presented in focus groups for discussion and validation. Interviewees willing to contribute further were invited to participate in focus groups [19] and refer other potential participants. Expert input across all One Health sectors was gathered through focus group discussions, which helped validate the relationships identified in interviews and identify additional variables or connections not captured initially.

Interview and focus group recordings were professionally transcribed and then analysed using MaxQDA. An inductive coding approach [20] was applied to identify themes relating to both direct and indirect factors driving AMR. Following thematic analysis, the five-step approach from Baugh Littlejohns, Baum [21] was adapted to determine the directionality (positive or negative) of relationships between variables and identify feedback loops.

The initial CLD was constructed in Stella Architect using coded interview data. It was subsequently presented during focus groups, where participants validated the variable names and relationships. The final diagram was refined and completed by the study team.

## 3. Results

A total of 35 participants from the human, animal and environmental health sectors took part in this study. This included 29 participants across 30 interviews (with one participant interviewed twice, before and after model development) and 10 participants in two focus group discussions. Interviews lasted approximately 60 minutes each, while each focus group ran for around 120 minutes.

Participants represented a diverse mix of professions, including clinicians (*n* = 15; e.g., infectious disease physicians, duty medical officers, rural medical practitioners, pharmacists and veterinarians); policymakers (*n* = 12; e.g., those working in health, housing, electricity and water services); researchers (*n* = 4); and others (*n* = 4; e.g., representatives from the private sector, including a pharmacy manager and a food systems consultant, and from a national policy advisory body). Participants were categorised into main One Health domains—human (*n* = 17), animal (*n* = 11), environmental and plant (*n* = 5), and cross-sectoral (*n* = 2)—as well as by professional background (Fig 1). Some participants have expertise spanning multiple domains or hold dual roles relevant to AMR. Specifically, five of the six physicians also have a background in chronic or infectious disease research; two veterinarians have experience in animal health research; and six policymakers in the animal health domain were also trained as veterinarians or researchers.

**Fig 1.**
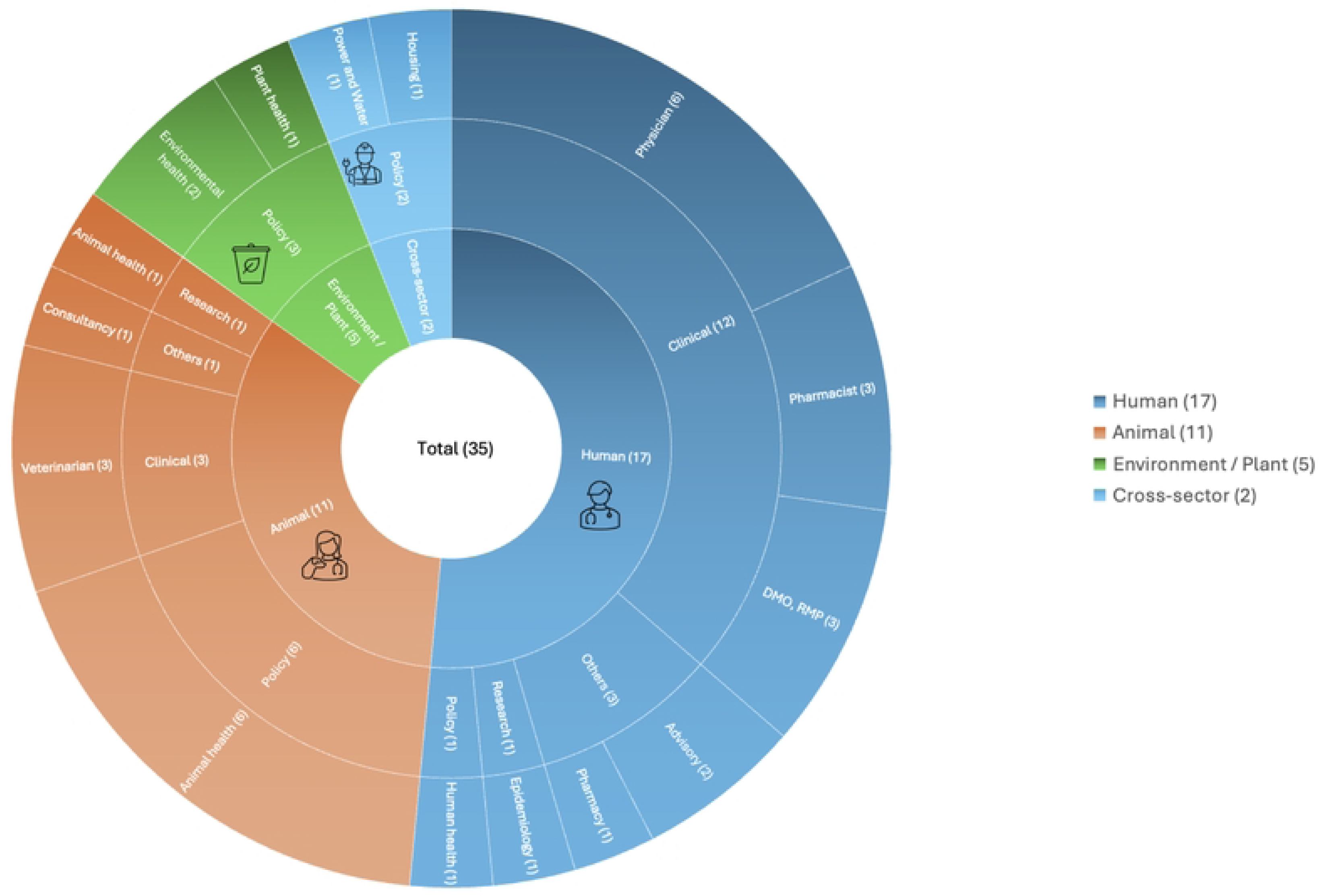
Study participants across One Health sectors (*n* = 35) DMO: duty medical officer; RMP: rural medical practitioner

Number in bracket indicates the number of participants in each One Health domain or professional category.

### 3.1. The One Health AMR model

Analysis of stakeholder input identified antibiotic prescribing and use in both human and animal health, as well as antibiotic residues in the environment, as key factors driving AMR evolution and spread in the NT (Fig 2). However, the dynamics underpinning AMR were found to extend beyond clinical settings, reflecting the broader complexity of the AMR system. A series of reinforcing and balancing feedback loops was identified, illustrating how interactions amongst factors influencing AMR can either amplify or mitigate its emergence and spread over time. The resulting One Health AMR model comprises four interconnected components—human healthcare, community, animal and environment—which are briefly described below.

**Fig 2.**
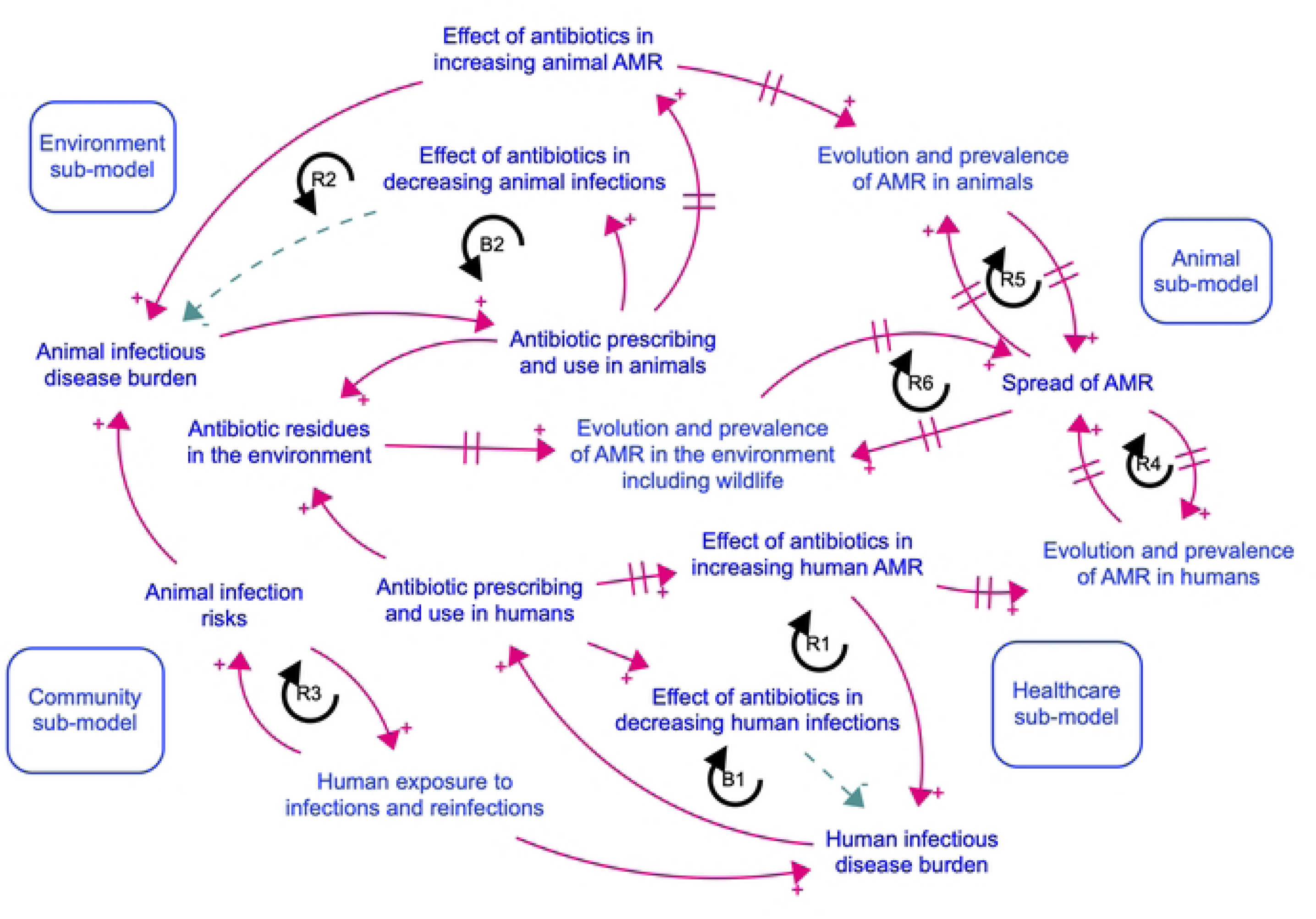
The overall causal loop model of AMR evolution and spread across One Health. 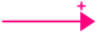: positive relationship; 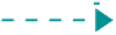: negative relationship; R and B: reinforcing and balancing loops; //: delay

The first component of the model focuses on how AMR may evolve and spread in humans. As illustrated in balancing loop B1 (Fig 2 and Table 1), antibiotic prescribing and use in humans is aimed at treating or preventing bacterial infections. A rise in infections increases the demand for antibiotics and subsequent prescribing (a ‘*positiv*e’ relationship). In turn, increased antibiotic use reduces infections, reflecting the intended therapeutic effect (a ‘*positive*’ relationship). As infections decrease due to effective treatment (a ‘*negative*’ relationship), the demand for antibiotics also declines. However, reinforcing loop R1 illustrates that increasing reliance on antibiotics to treat infections contributes to the emergence of AMR (a ‘*positiv*e’ relationship), which subsequently results in treatment failures and a rise in infections (a ‘*positive*’ relationship). The development of AMR is often delayed (indicated by ‘//’) as resistance typically emerges sometime after initial exposure to antibiotics rather than occurring immediately [22]. This delay can contribute to an underestimation of the long-term consequences of AMR, reinforcing the cycle of escalating infections and greater antibiotic demand. The evolution and spread of AMR in human health is shaped by a range of direct and indirect factors, which are further examined in Section 3.2 (healthcare sub-model) and 3.3 (community sub-model).

**Table 1.** Structure of feedback loops in the One Health AMR model.

| **Loop number** | **Loop structure** |
| --- | --- |
| *Reinforcing loops* |  |
| R1 | Antibiotic prescribing and use in humans →(+) Effect of antibiotics in increasing human AMR →(+) Human infectious disease burden →(+) Antibiotic prescribing and use in humans |
| R2 | Antibiotic prescribing and use in animals →(+) Effect of antibiotics in increasing animal AMR →(+) Animal infectious disease burden →(+) Antibiotic prescribing and use in animals |
| R3 | Human exposure to infections and reinfections →(+) Animal infection risks →(+) Human exposure to infections and reinfections |
| R4 | Evolution and prevalence of AMR in humans →(+) Spread of AMR →(+) Evolution and prevalence of AMR in humans |
| R5 | Evolution and prevalence of AMR in animals →(+) Spread of AMR →(+) Evolution and prevalence of AMR in animals |
| R6 | Evolution and prevalence of AMR in the environment including wildlife →(+) Spread of AMR →(+) Evolution and prevalence of AMR in the environment including wildlife |
| *Balancing loops* |  |
| B1 | Antibiotic prescribing and use in humans →(+) Effect of antibiotics in decreasing human infections →(-) Human infectious disease burden →(+) Antibiotic prescribing and use in humans |
| B2 | Antibiotic prescribing and use in animals →(+) Effect of antibiotics in decreasing animal infections →(-) Animal infectious disease burden →(+) Antibiotic prescribing and use in animals |

→(+) and →(-): positive and negative relationships; R and B: reinforcing and balancing loops

The second component of the model focuses on the emergence and spread of AMR within animal populations and their interconnection with human health. As illustrated in loops B2 and R2, AMR evolution in animals is similarly driven by antibiotic use. The risk of AMR spread is bi-directional, whereby infections in animals can increase human exposure and vice versa (loop R3). Similar reinforcing feedback mechanisms drive the evolution and spread of AMR in both animal and human hosts (loops R4-5).

Lastly, in the environmental domain, antibiotic residues originating from both human and animal health systems were identified as a key contributor to AMR. These residues may exert selective pressure driving the evolution of resistant organisms, while wildlife and the broader environmental sources facilitate its spread. Resistant microbes or resistant genes may re-enter human and animal systems via pathways such as water, air, soil or direct contact [23, 24], thus amplifying the One Health transmission cycle (loops R4-6).

To enhance clarity and focus on the feedback loops critical to system behaviour, the CLD was divided into sub-models. These sub-models were informed by themes emerging from interview data that shaped the identification of factors driving AMR. This paper discusses in detail the interconnected factors influencing AMR in human health, as captured in the healthcare (Section 3.2) and community (Section 3.3) sub-models. Details of each relationship and feedback loops are provided in S1-4 Files.

### 3.2. The healthcare sub-model

The human healthcare sub-model reflects participant perspectives on factors driving AMR and their interactions across both hospital and primary healthcare settings in the NT. It consists of 56 factors and 89 connections (Fig 3 and S1 File), generating 118 reinforcing and 101 balancing feedback loops (S2 File). Reinforcing loop R1 (Fig 3 or Table 2) illustrates a feedback mechanism in which increased reliance on antibiotics for treating infections elevates selective pressure, thereby heightening the AMR risk. This, in turn, contributes to treatment failures, rising infection incidence, and continued antibiotic demand, amplifying the risk of medication-related adverse events. One clinician reported scenarios where first-line antimicrobials failed due to resistance, necessitating the use of last-resort agents such as colistin to manage severe infections (loop B1). While effective in some instances, such antibiotics are often associated with serious side effects (S1 Table, quote 1.1). In certain cases, this may be nephrotoxic, resulting in acute kidney injury [25] and requiring interventions such as dialysis (loop B3).

**Fig 3.**
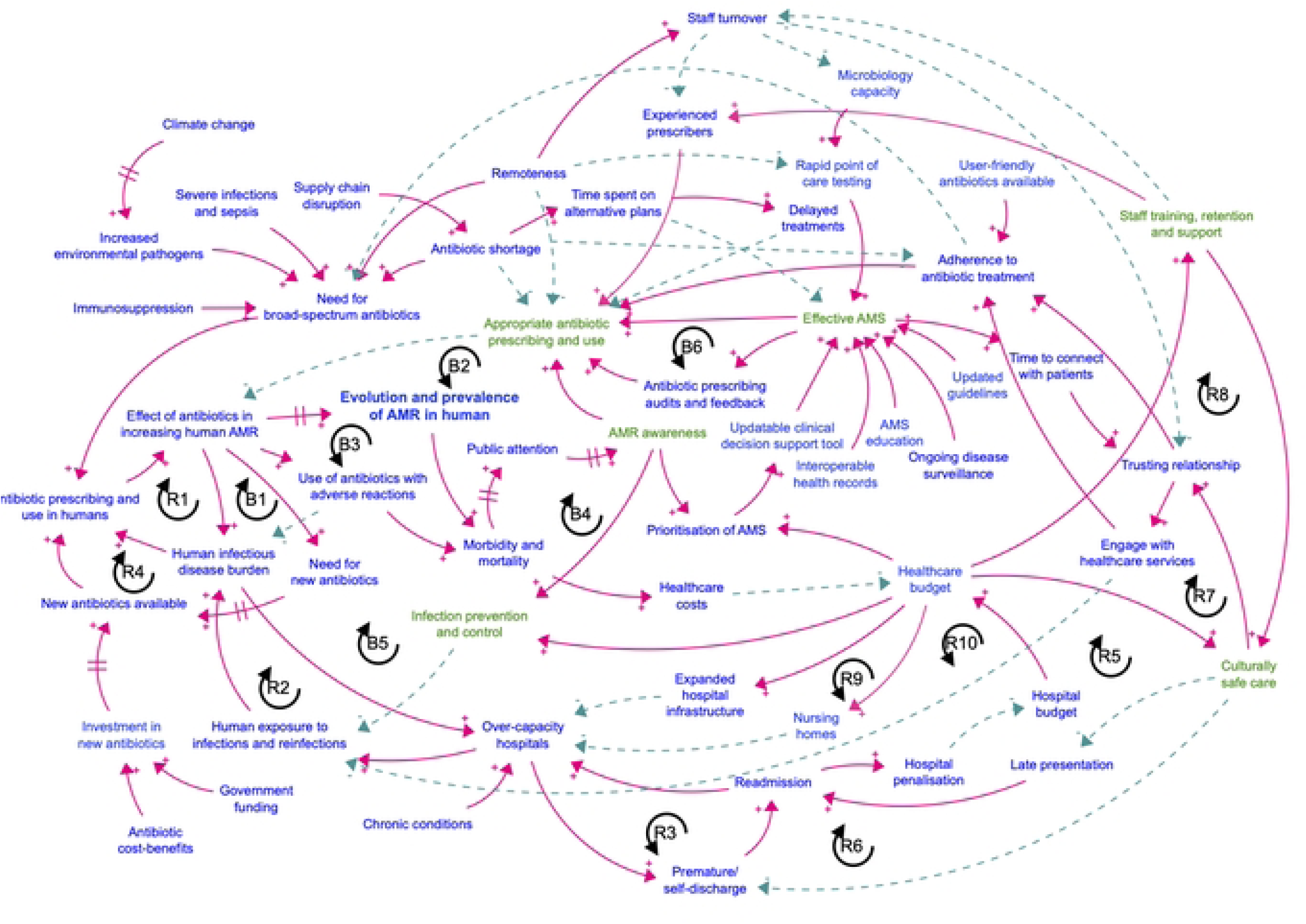
AMR evolution and spread in the human healthcare sub-model. 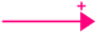: positive relationship; 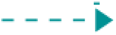: negative relationship; R and B: reinforcing and balancing loops; //: delay; Green colour: leverage points for interventions to address AMR.

**Table 2.**
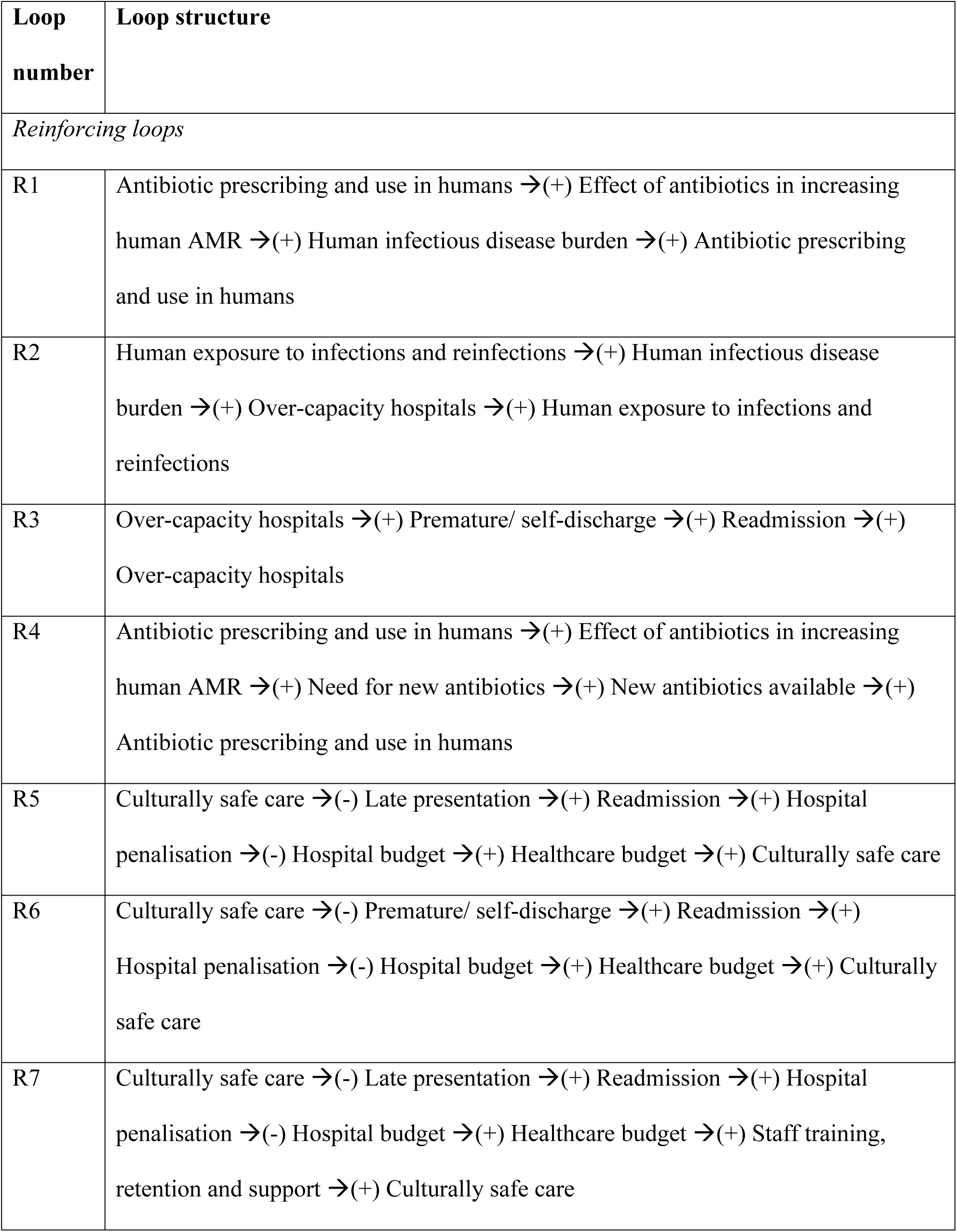
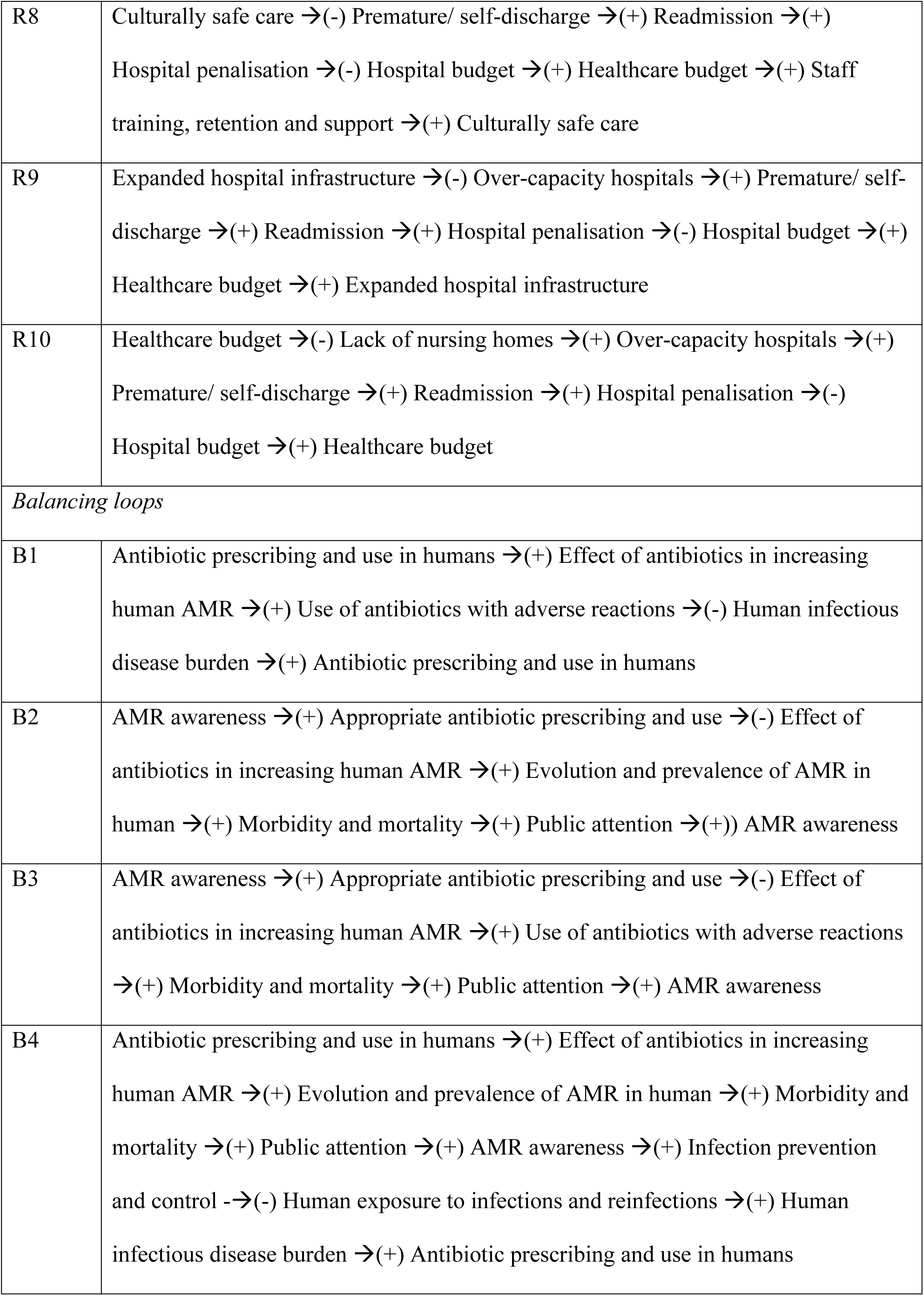
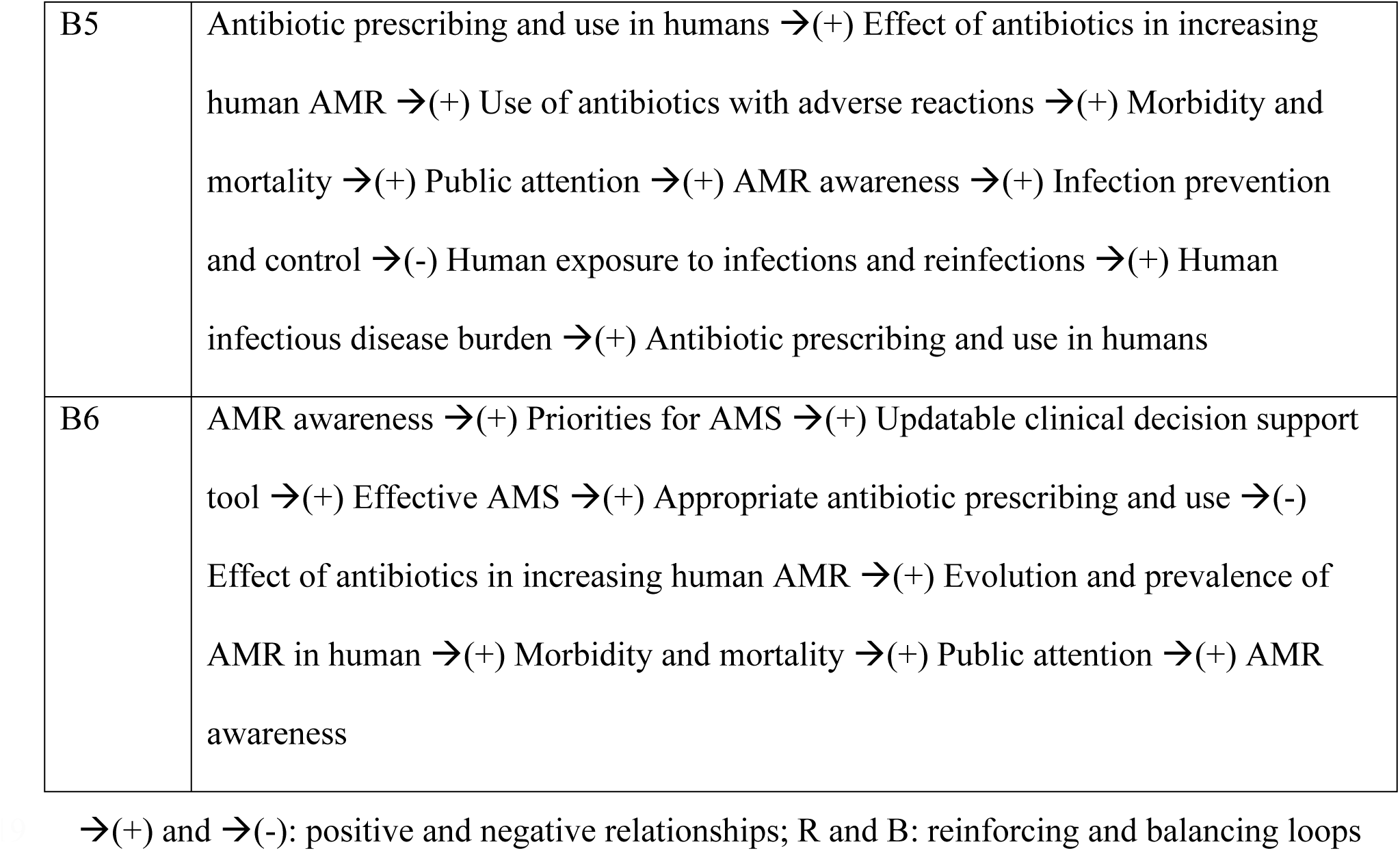
Key feedback loops in the human health sub-model.

| **Loop number** | **Loop structure** |
| --- | --- |
| *Reinforcing loops* |  |
| R1 | Antibiotic prescribing and use in humans →(+) Effect of antibiotics in increasing human AMR →(+) Human infectious disease burden →(+) Antibiotic prescribing and use in humans |
| R2 | Human exposure to infections and reinfections →(+) Human infectious disease burden →(+) Over-capacity hospitals →(+) Human exposure to infections and reinfections |
| R3 | Over-capacity hospitals →(+) Premature/ self-discharge →(+) Readmission →(+) Over-capacity hospitals |
| R4 | Antibiotic prescribing and use in humans →(+) Effect of antibiotics in increasing human AMR →(+) Need for new antibiotics →(+) New antibiotics available →(+) Antibiotic prescribing and use in humans |
| R5 | Culturally safe care →(-) Late presentation →(+) Readmission →(+) Hospital penalisation →(-) Hospital budget →(+) Healthcare budget →(+) Culturally safe care |
| R6 | Culturally safe care →(-) Premature/ self-discharge →(+) Readmission →(+) Hospital penalisation →(-) Hospital budget →(+) Healthcare budget →(+) Culturally safe care |
| R7 | Culturally safe care →(-) Late presentation →(+) Readmission →(+) Hospital penalisation →(-) Hospital budget →(+) Healthcare budget →(+) Staff training, retention and support →(+) Culturally safe care |
| R8 | Culturally safe care →(-) Premature/ self-discharge →(+) Readmission →(+)<br>Hospital penalisation →(-) Hospital budget →(+) Healthcare budget →(+) Staff training, retention and support →(+) Culturally safe care |
| R9 | Expanded hospital infrastructure →(-) Over-capacity hospitals →(+) Premature/ self-discharge →(+) Readmission →(+) Hospital penalisation →(-) Hospital budget →(+) Healthcare budget →(+) Expanded hospital infrastructure |
| R10 | Healthcare budget →(-) Lack of nursing homes →(+) Over-capacity hospitals →(+) Premature/ self-discharge →(+) Readmission →(+) Hospital penalisation →(-) Hospital budget →(+) Healthcare budget |
| *Balancing loops* |  |
| B1 | Antibiotic prescribing and use in humans →(+) Effect of antibiotics in increasing human AMR →(+) Use of antibiotics with adverse reactions →(-) Human infectious disease burden →(+) Antibiotic prescribing and use in humans |
| B2 | AMR awareness →(+) Appropriate antibiotic prescribing and use →(-) Effect of antibiotics in increasing human AMR →(+) Evolution and prevalence of AMR in human →(+) Morbidity and mortality →(+) Public attention →(+) AMR awareness |
| B3 | AMR awareness →(+) Appropriate antibiotic prescribing and use →(-) Effect of antibiotics in increasing human AMR →(+) Use of antibiotics with adverse reactions →(+) Morbidity and mortality →(+) Public attention →(+) AMR awareness |
| B4 | Antibiotic prescribing and use in humans →(+) Effect of antibiotics in increasing human AMR →(+) Evolution and prevalence of AMR in human →(+) Morbidity and mortality →(+) Public attention →(+) AMR awareness →(+) Infection prevention and control →(-) Human exposure to infections and reinfections →(+) Human infectious disease burden →(+) Antibiotic prescribing and use in humans |
| B5 | Antibiotic prescribing and use in humans →(+) Effect of antibiotics in increasing human AMR →(+) Use of antibiotics with adverse reactions →(+) Morbidity and mortality →(+) Public attention →(+) AMR awareness →(+) Infection prevention and control →(-) Human exposure to infections and reinfections →(+) Human infectious disease burden →(+) Antibiotic prescribing and use in humans |
| B6 | AMR awareness →(+) Priorities for AMS →(+) Updatable clinical decision support tool →(+) Effective AMS →(+) Appropriate antibiotic prescribing and use →(-) Effect of antibiotics in increasing human AMR →(+) Evolution and prevalence of AMR in human →(+) Morbidity and mortality →(+) Public attention →(+) AMR awareness |

→(+) and →(-): positive and negative relationships; R and B: reinforcing and balancing loops

Key themes emerged from interviews and focus groups include human antibiotic prescribing and use, patient adherence to antibiotic treatment, antimicrobial stewardship, and healthcare infrastructure and culturally safe care.

#### Antibiotic prescribing and use

High antibiotic use in the NT is driven by many interrelated clinical and structural factors. The high burden of severe infections and sepsis, compounded by significant prevalence of chronic conditions such as diabetes, kidney and cardiovascular disease [26, 27] predisposes individuals in this region to recurrent infections.

Broad-spectrum antibiotics are commonly used to manage or prevent secondary bacterial infections, such as those resulting from viral respiratory infections and scabies-related skin infections, and to reduce sepsis-related deaths. This practice may, in part, reflect a preventive strategy aimed at reducing the risk of complications, including rheumatic heart disease (S1 Table, quotes 1.2-1.4).

Additionally, the increasing frequency of extreme weather events and climate variability in the NT may contribute to the incidence and spread of environmental pathogens, including *Burkholderia pseudomallei* and *Vibrio* spp., posing significant infection risks—such as melioidosis [28, 29]—particularly for those with underlying health conditions (S1 Table, quote 1.5). This persistent burden of both chronic and infectious diseases necessitates frequent use of broad-spectrum antibiotics. Prophylactic use is also common in remote areas due to limited diagnostic capacity, shortages of narrow-spectrum antibiotics, and issues with adherence to treatment (S1 Table, quotes 1.6-1.8).

#### Patient adherence to antibiotic treatment

Patient adherence to treatment is influenced by access to and engagement with healthcare services, both of which are underpinned by trusting relationships (S1 Table, quotes 2.1-2.2) between patients and healthcare providers (loops R22-26, S2 File). In the NT, high staff turnover, driven by geographic isolation, has led to increased reliance on fly-in fly-out healthcare workers, making it difficult to establish trust and continuity of care [30].

Adherence can be improved by the availability of ‘user-friendly’ antibiotic formulations, such as easy-to-swallow tablets or medications requiring less frequent dosing (S1 Table, quote 2.3). Adherence, in turn, can also influence antibiotic prescribing choices (loops R15-16, R22-25, S2 File), potentially compromising antimicrobial stewardship if ease of administration is prioritised over targeted treatment, thereby increasing AMR risk.

#### Antimicrobial stewardship

Antimicrobial stewardship (AMS) programs aim to optimise antimicrobial use, improve patient outcomes and reduce AMR risk. In the NT, limitations in AMS [31] are largely due to inadequate prioritisation, potentially stemming from low awareness of AMR severity (loop B6, Table 2) and constrained healthcare budget (loops B18 and B20, S2 File). Clinicians highlighted that incorporating timely, local AMR data from disease surveillance systems and point-of-care testing is critical to guide effective AMS. Additionally, access to up-to-date treatment guidelines, integrated AMS education, and interoperable electronic health records across hospital and primary care are essential. A critical missing element is a contemporary, updatable clinical decision support tool (S1 Table, quotes 3.1-3.2).

#### Healthcare infrastructure and culturally safe care

Participants also identified systemic healthcare factors contributing AMR. Overcrowded hospitals, due to the high burden of infections and chronic diseases, long wait times for aged care admission, along with ageing infrastructure and limited single-patient rooms all contribute to increasing infections and reinfections (loops R2 and R10, Table 2) and AMR (S1 Table, quotes 4.1-4.3).

A high self-discharge rate from NT hospitals [32, 33], partly due to cultural safety issues, poor communication, extended wait times or family responsibilities, can elevate readmissions, exacerbate hospital strain (loops R3 and R5) and impact healthcare budgets (loops R9-10). Premature or self-discharge can subsequently reduce hospital revenue, which is often tied to inpatient days, further limiting the capacity to deliver culturally safe care (loop R6) (S1 Table, quotes 4.4-4.7).

A lack of focus on staff training, retention and support may also hinder culturally safe care. Loops R7 and R8 describe how these deficits contribute to late presentations, repeat readmissions and escalating healthcare costs (S1 Table, quotes 4.8-4.10).

To improve culturally safe care, participants emphasised the need for targeted staff training, especially comprehensive orientation for healthcare staff, including fly-in fly-out workers, to improve understanding of the NT context, alongside robust staff retention and support programs to reduce turnover (S1 Table, quotes 4.11-4.13). These, in turn, would foster trust, enhance patient engagement and improve adherence to antibiotic treatment.

#### Key leverage points

Numerous reinforcing loops illustrate how antibiotic prescribing and use are driven by interconnected factors (Fig 3 or S2 File), contributing to the evolution and spread of AMR in the NT. Yet, key leverage points—highlighted in green in the CLD—were identified as opportunities to reverse this dominant trend of AMR. These include (1) rational antibiotic prescribing and use (loops B2-3), which can be improved through (2) implementing effective AMS programs (loop B6), (3) raising awareness of antibiotic use and AMR (among both healthcare professionals and the community, (4) improving culturally safe care, and (5) investing in staff training, retention and support (loops R7-8). There is also a need to (6) strengthening infection prevention and control (loops B4-5).

Critically, clinical participants emphasised the need to reduce the high burden of infectious diseases as a strategy to ease pressure on the healthcare system. They highlighted the importance of preventive efforts at the community level to stop infections before they occur. These upstream strategies are detailed further in the community sub-model, which outlines structural factors identified by participants as essential for reducing the risk of AMR and infectious diseases in the NT.

### 3.3. The community sub-model

The community sub-model characterises the dynamics of AMR evolution and spread in non-healthcare settings. With overlapping in parts with the healthcare sub-model, it highlights the multidirectional pathways of AMR transmission between these two settings (Fig 4). The sub-model contains 43 variables and 79 connections (Fig 4 and S3 File), forming 40 reinforcing and one balancing feedback loops (S4 File).

**Fig 4.**
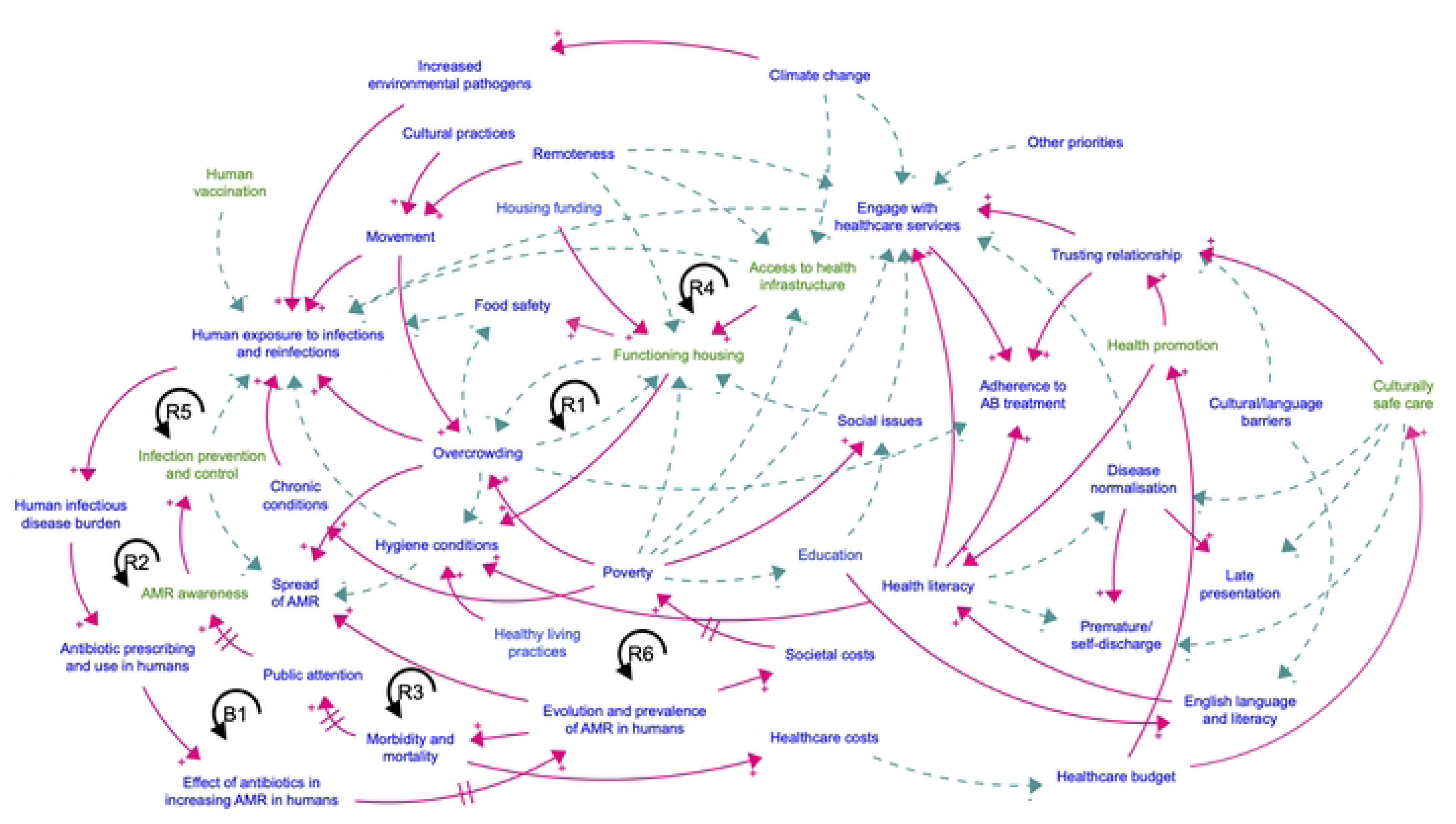
AMR evolution and spread in the community sub-model. 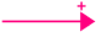: positive relationship; 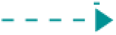: negative relationship; R and B: reinforcing and balancing loops; //: delay; Green colour: leverage points for interventions to address AMR.

Key themes emerging from interviews and focus groups include overcrowding and housing infrastructure, access to health hardware, and engagement with healthcare services.

#### Overcrowding and housing infrastructure

Participants identified household overcrowding as a critical structural determinant influencing both limited access to functional housing and increased exposure to infectious disease. This, in turn, contributes to recurrent infections and elevated AMR risk.

Overcrowded and poorly functioning housing, often compounded by poverty, significantly impact hygiene and food security, increasing susceptibility to infections [34] (S2 Table, quotes 5.1-5.3).

The existing housing shortage contributes to high household occupancy rates, with multiple individuals routinely sharing living and sleeping space. Housing design often fails to accommodate cultural practices and living norms of remote-dwelling populations. Under overcrowded conditions, housing functionality is further reduced (loop R1, Fig 4 or Table 3), as essential amenities often come under strain. These issues are particularly severe in remote areas, where poverty and social challenges are more widespread (S2 Table, quotes 5.4-5.7).

**Table 3.** Key feedback loops in the community sub-model.

| Loop number | Loop structure |
| --- | --- |
| *Reinforcing loops* |  |
| R1 | Overcrowding →(-) Functioning housing →(-) Overcrowding |
| R2 | Access to health infrastructure →(-) Human exposure to infections and reinfections →(+) Human infectious disease burden →(+) Antibiotic prescribing and use in humans →(+) Effect of antibiotics in increasing AMR in humans →(+) Evolution and prevalence of AMR in humans →(+) Societal costs →(+) Poverty →(-) Access to health infrastructure |
| R3 | Antibiotic prescribing and use in humans →(+) Effect of antibiotics in increasing AMR in humans →(+) Evolution and prevalence of AMR in humans →(+) Societal costs →(+) Poverty →(+) Chronic conditions →(+) Human exposure to infections and reinfections →(+) Human infectious disease burden →(+) Antibiotic prescribing and use in humans |
| R4 | Antibiotic prescribing and use in humans →(+) Effect of antibiotics in increasing AMR in humans →(+) Evolution and prevalence of AMR in humans →(+) Societal costs →(+) Poverty →(-) Engage with healthcare services →(-) Human exposure to infections and reinfections →(+) Human infectious disease burden →(+) Antibiotic prescribing and use in humans |
| R5 | Antibiotic prescribing and use in humans →(+) Effect of antibiotics in increasing AMR in humans →(+) Evolution and prevalence of AMR in humans →(+) Societal costs →(+) Poverty →(+) Overcrowding →(+) Human exposure to infections and reinfections →(+) Human infectious disease burden →(+) Antibiotic prescribing and use in humans |
| R6 | Antibiotic prescribing and use in humans →(+) Effect of antibiotics in increasing AMR in humans →(+) Evolution and prevalence of AMR in humans →(+) Societal costs →(+) Poverty →(-) Functioning housing →(+) Food safety →(-) Human exposure to infections and reinfections →(+) Human infectious disease burden →(+) Antibiotic prescribing and use in humans |

| *Balancing loops* |  |
| --- | --- |
| B1* | AMR awareness →(+) Infection prevention and control →(-) Human exposure to infections and reinfections →(+) Human infectious disease burden →(+) Antibiotic prescribing and use in humans →(+) Effect of antibiotics in increasing AMR in humans →(+) Evolution and prevalence of AMR in humans →(+) Morbidity and mortality →(+) Public attention →(+) AMR awareness |

\* Similar to loop B4 in the healthcare sub-model
→(+) and →(-): positive and negative relationships; R and B: reinforcing and balancing loops

While timely maintenance of housing infrastructure remains a challenge in many remote communities, programs such as *Healthy Homes* (Grealy et al., 2025) represent important steps toward building local capacity for housing repair and maintenance. Participants acknowledged the value of such initiatives in addressing long-standing infrastructure issues but also noted challenges stemming from competing priorities and limited funding (S2 Table, quote 5.8).

#### Access to health hardware

In addition to housing availability and quality, the accessibility of essential health-supporting infrastructure, including safe water and reliable electricity, remains a critical determinant of health in the NT. While bore water is the primary water source in many remote communities, certain areas have reported chemical or physical exceedances of Australian drinking water guidelines. The increasingly hot climate and extreme weather conditions in the NT complicate the maintenance and durability of water infrastructure (S2 Table, quotes 6.1-6.2). As a result, this degradation increases the likelihood of microbial contamination and chemical pollutants, compromising the safety and quality of water supplied to communities.

Poor water quality and inadequate access to basic utilities, combined with overcrowding, likely contribute to high rates of skin and soft tissue infections. Such infections, if left untreated, can progress to severe sequelae, such as sepsis, rheumatic fever and rheumatic heart disease, which is highly prevalent in remote communities [35, 36].

#### Engagement with healthcare services

Participants also emphasised a range of factors contributing to low engagement with healthcare services. While geographic isolation presents a physical barrier, individuals may remain reluctant to seek care even when services are accessible. Reflecting patterns identified in the healthcare sub-model, this reluctance is often due to complex drivers such as limited trust in the healthcare system and the absence of culturally safe care (S2 Table, quotes 7.1-7.3)

Additional contributing factors include cultural and language barriers, low health literacy and the normalisation of disease, which together reduce the perceived urgency of seeking medical care. Limited health literacy can impair individuals’ ability to understand, navigate and effectively engage with health services, thereby reducing overall uptake and health-seeking behaviour. The normalisation of conditions such as impetigo and scabies infestation in remote communities [37], often compounded by stigma, can reduce willingness to seek care and delay recognition of illness severity by both patients and healthcare providers. Other reasons for delayed care-seeking include competing priorities such as family or carer responsibilities or ‘sorry business’ (S2 Table, quotes 7.4-7.9).

These factors limit engagement with healthcare services and delay initiation of appropriate treatment. The interplay between low health literacy and disease normalisation may also contribute to high rates of self-discharge from healthcare facilities.

#### Key leverage points

The reinforcing loops shown in Fig 4 or S4 File illustrate how various factors interact to drive infections and recurrent infections in the community. The societal costs related to AMR (e.g., prolonged hospital stays and productivity losses) further exacerbate persistent challenges such as poverty, overcrowding, and chronic conditions (loops R2-R6). Breaking this vicious cycle requires addressing key leverage points identified in the model, including (1) increasing access to functional housing and utilities (loop R1), (2) enhancing culturally safe care (loops R25-26, S4 File) and (3) continuing community-based health promotion (loops R27-29, S4 File), among other factors discussed in Section 3.2. This close integration of community and clinical environments highlights their interdependence and the importance of coordinated action to reduce AMR transmission within and across both sub-systems.

## 4. Discussion

Drawing on input from stakeholders across all One Health sectors, we developed a causal loop model to conceptualise the direct and indirect factors driving the evolution and spread of AMR in a high burden region of Australia. By focusing on regional and remote settings, our findings demonstrate how the complex interrelationships among factors driving AMR shape the dynamics of infectious diseases and resistance. These findings highlight that addressing AMR through isolated interventions is insufficient and unlikely to result in sustained reductions in disease burden; thus, a systems-based approach is critical. For example, despite widespread antibiotic use, infections such as trachoma and recurrent scabies persist in the region (S1 Table, quote 1.9), reflecting a broader failure to address upstream or primordial determinants of health.

We found that the prevailing narrative attributing the high burden of AMR in the NT to high volumes of broad-spectrum antibiotic use needs critical re-evaluation. Given the disproportionately high burden of infectious diseases, the risk of AMR is compounded by systemic challenges, including healthcare delivery and access in geographically remote areas with high staff turnover [38], substandard housing infrastructure [34], and socio-economic disadvantage [39]. In this context, the use for antibiotics, including new agents, is both inevitable and essential to reduce morbidity and mortality. What may be externally perceived as antibiotic overuse is, in fact, often a clinically appropriate and necessary response to prevent serious infections and adverse outcomes. This aligns with previous research that reported high levels of appropriate antibiotic prescribing following standard guidelines (over 85%) across regional and remote primary care settings of northern Australia, including the NT [40]. Moreover, what remains underexplored is the role of resistant strains and resistance genes in both healthcare and non-healthcare environments as dominant contributors to AMR. Consequently, reducing antibiotic use alone is unlikely to sufficiently control AMR [41]. While the development of new antibiotics is important, reliance on pharmaceutical innovation alone will continue to perpetuate a cycle in which resistance eventually re-emerges. Without addressing the underlying systemic drivers of AMR, such advances are likely to provide only temporary relief.

Our approach identified key leverage points for high-impact interventions that may produce sustainable change and minimise unintended consequences across both healthcare and community settings. These include interoperable AMS and infection control programs, culturally safe care, and healthcare workforce training, retention and support across different healthcare settings; AMR awareness; improved access to functional housing and health infrastructure; and community-based health promotion.

As illustrated in our model, while new antibiotics may offer short-term relief, they target only the symptoms of AMR rather than its root causes. Since the introduction of new vaccines depends on external factors, other preventive measures, particularly enhanced infection prevention and control, are more likely to offer sustainable solutions by reducing infectious disease burden and subsequent demand for antibiotics. Participants consistently emphasised the importance of addressing infections at their source as a key strategy to mitigate AMR risk. This requires a shift in focus from reactive, clinical responses to proactive, community-based interventions.

Continued investment in functional housing, community-led maintenance programs, and access to health hardware is essential to reduce overcrowding and its associated health impacts. Improvements in cultural safety and continuity of care, including more opportunities for staff training, retention and support [42], as well as effective communication [33], alongside AMR awareness and health promotion, are critical for building trust, enhancing healthcare engagement and promoting treatment adherence. Though such investments require resources, they have the potential to significantly reduce the burden of infections and AMR, thereby alleviating pressures on both the healthcare system and society at large.

## Limitations

The CLD developed in this study illustrated key interconnected factors influencing AMR in humans, providing a comprehensive systems-level view. However, the model does not capture all relationships—a limitation that reflects both the scope of this study and the inherent complexity of AMR. For example, community voices and the lived experiences of patients have not yet been incorporated. While input from service providers in health, housing and utilities provided valuable insights at the population level, future research would benefit from community engagement to elicit individual-level perspectives.

Another limitation lies in the challenge of prioritising the many pressing issues associated with AMR in the NT. While stakeholders identified a wide range of needs, we focused on leverage points highlighted during focus group discussions. These represent key opportunities to shift the trajectory of the system, particularly by reducing infection burden and long-term AMR risk. We acknowledge that biases may exist, and our ongoing work aims to test our hypotheses using quantitative system dynamics simulation based on real-world data. This next phase will allow further refinement of the model and inform the design and evaluation of effective policy and management strategies.

## 5. Conclusions

This study built on stakeholder perspectives to explore the interrelated factors driving the evolution and spread of AMR with a focus on human health. These insights were synthesised into a visual model—a causal loop diagram—that represents our hypothesis of the key contributors to the problem. While many of the issues described have been identified in previous studies, our model highlights how these factors are interconnected and collectively contribute to the dynamic complexity of the AMR system. The causal loop model is a valuable tool for uncovering root causes and hidden drivers and for capturing a holistic picture of the infectious disease and AMR burden in the NT. Importantly, this approach enables stakeholders to adopt a systems thinking mindset to explore the behaviour of complex systems such as AMR, and provides a platform to identify opportunities for targeted intervention strategies that can inform more effective decision-making. The causal loop model remains a work in progress and future iterations will incorporate richer discussions of currently missing elements, particularly community voices and lived experiences.

As part of the broader project, we are also investigating the direct and indirect factors driving AMR in animal and environmental systems, including wildlife, and their interactions with human health. Understanding the complex influences in shaping AMR dynamics within these reservoirs is critical, yet remains underrepresented in AMR research and surveillance efforts [43–45].

The next phase of our work involves the development of a system dynamics simulation model. This will allow us to design and test alternative policy and practice scenarios and quantify their potential effectiveness in reducing AMR risk, thereby informing evidence-based decision-making.

Overall, our findings reinforce that addressing AMR cannot rely solely on a single system, particularly healthcare system interventions or antimicrobial stewardship in isolation. In the context of the NT, achieving sustained reductions in AMR requires addressing the primordial determinant of health and shifting towards proactive, community-based interventions. There is no ‘silver-bullet’ to tackle the multifaceted, systemic challenges of AMR. Meaningful and sustainable progress will require a coordinated, multi-sectoral, and systems-informed approach that integrates perspectives across health, environmental and community domains.

## Data Availability

All relevant data are available through anonymised quotations provided in the Supporting Information files, in accordance with the requirements of the relevant Ethics Committees.

## Acknowledgements

This work was supported by the CSIRO Early Research Career Postdoctoral Fellowship awarded to the first author. The authors gratefully acknowledge the valuable contributions of all study participants. We extend special thanks to Professor Bart Currie and Dr Jessica Hoopes for their insightful advice and feedback on the draft manuscript.

## Supporting information

**S1 File. Relationships between factors in the healthcare sub-model**

**S2 File. Feedback loops in the healthcare sub-model**

**S3 File. Relationships between factors in the community sub-model**

**S4 File. Feedback loops in the community sub-model**

**S1 Table. Themes and quotes relating to factors driving antimicrobial resistance in the healthcare sub-model**

**S2 Table. Themes and quotes relating to factors driving antimicrobial resistance in the community sub-model**

